# Strengthening Cardiac Rehabilitation: Key Strategies for Enhancing Accessibility and Outcomes

**DOI:** 10.64898/2026.08.20.26360821

**Authors:** Raghad Tawalbeh, Jullie Ellis, Kyle T. Ebersole, Kim Litwack

**Author notes:** **Corresponding Author:** Raghad Tawalbeh, PhD, MSN, RN^1,2^, School of Nursing, University of North Carolina Greensboro, 1000 Spring Garden St, Greensboro, NC 27412, USA. Associate Professor, University of Wisconsin–Milwaukee, School of Nursing, 3450 N. Downer Avenue, Milwaukee, WI 53211, USA. Professor and Director, Human Performance & Sport Physiology Laboratory, University of Wisconsin–Milwaukee, School of Rehabilitation Sciences & Technology, 3200 N. Cramer Street, Milwaukee, WI 53211, USA. Professor and Dean, College of Health Professionals, University of Wisconsin–Milwaukee, School of Nursing, 3450 N. Downer Avenue, Milwaukee, WI 53211, USA.

## Abstract

**Introduction:** Cardiac rehabilitation (CR) is key for secondary prevention; however, participation remains low due to persistent barriers. Identifying strategies used by high-performing programs may inform approaches to improve patient engagement and outcomes.

**Purpose:** To identify strategies associated with improved participation and adherence in CR programs from the perspective of leaders in high-performing sites.

**Methods:** Semi-structured interviews were conducted with 10 CR leaders from urban, suburban, and rural programs ranked in the top 10% on at least two objective performance measures (e.g., participation and adherence rates) but moderate or low on others. Data were analyzed using thematic analysis to identify strategies associated with high performance.

**Results:** Programs with high participation and adherence rates consistently implemented proactive, patient-centered strategies to address barriers. Individualized care approaches tailored to language, culture, health literacy, and age were commonly used to improve engagement among Hispanic, Black, and older adult populations. High-performing programs addressed structural barriers such as insurance and transportation through flexible scheduling, community partnerships, and targeted outreach. Strong coordination with referring providers and effective transitions from inpatient to outpatient care were associated with higher enrollment and sustained participation. Additional strategies included staff development through ongoing education, use of digital tools for patient tracking, and implementation of virtual and hybrid CR models. Integration of psychological support further enhanced patient engagement.

**Conclusion:** High-performing CR programs employ coordinated, patient-centered, and system-level strategies associated with improved participation and adherence. These findings provide actionable approaches to enhance accessibility and improve programs and patients outcomes in CR across diverse settings.

## Introduction

Cardiac rehabilitation is a multidisciplinary program designed to improve overall health and well-being after a cardiac event ^1–5^. It includes exercise training, education on healthy lifestyle habits, stress management, and support for psychological well-being.^1,2,4,6^. CR is delivered through center-based (CBCR), home-based (HBCR), and hybrid models that promote physical activity, healthy behaviors, medication adherence, and psychosocial well-being to improve cardiovascular outcomes. ^7,8^.

The American Association of Cardiovascular and Pulmonary Rehabilitation (AACVPR) developed a 2022 cardiac rehabilitation (CR) measure set that includes nine measures, six performance and three quality measures, applicable to both inpatient and outpatient settings. Key performance areas include functional capacity, blood pressure, smoking cessation, enrollment, and adherence, all aimed at reducing cardiovascular risk factors and improving overall health.^2,5,9^

Despite the well-established benefits of CR and the availability of multiple delivery models, participation rates remain consistently low, with many eligible patients never enrolling or failing to complete programs ^10,11^. Barriers related to accessibility, including geographic limitations, transportation challenges, socioeconomic factors, and limited program availability—continue to restrict equitable access to CR services ^12,13^. Furthermore, variability in program delivery and implementation of established guidelines may contribute to inconsistent patient engagement and outcomes ^14–17^. Addressing these gaps requires a deeper understanding of how CR leaders navigate these challenges and implement strategies to improve access, participation, and overall program effectiveness.

### Study purpose

This study aims to identify strategies associated with improved participation and adherence in CR programs from the perspective of leaders in high-performing sites.

## Methods

We used a qualitative descriptive design with semi-structured interviews to explore CR leaders’ perspectives on strategies to improve program accessibility and outcomes, including participation, adherence, and patient-centered care. Semi-structured interviews allowed in-depth exploration of internal program processes and decision-making.

### Sampling

Purposive sampling of participants from CR programs ranked in the top 10% based on at least one AACVPR performance measure. CR leaders were eligible if they held decision-making roles related to program structure and delivery. A total of 10 participants included, with sampling continuing until achieving sufficient depth of information.

### Data Collection

The interview guide aimed to explore challenges, gaps, and strategies related to CR implementation and accessibility. We conducted the interviews virtually, lasted 30–80 minutes, audio-recorded, transcribed verbatim, and securely stored.

### Bias Reduction

Although performance data were available through the AACVPR registry, neither participants nor the interviewer were informed of specific performance measures. However, program leaders were able to independently access their own program information if they chose to do so. This approach helped reduce response bias and encouraged broader reflection on program practices.

### Data Analysis

We conducted deductive content analysis using a predefined coding framework derived from existing literature and study objectives. We applied constant comparison to identify patterns and refine themes, strengthening analytic rigor and consistency.

## Results

Across interviews with CR leaders, four interrelated themes emerged: (1) multi-level barriers to CR participation, (2) equity implications of these barriers, (3) adaptive patient-centered and programmatic strategies to improve access and outcomes, and (4) system-level innovations and future directions. Together, these themes illustrate how CR programs are simultaneously constrained by structural and patient-level challenges while actively implementing strategies to improve participation, adherence, and equity in care delivery.

### Challenges and gaps in CR implementation

The CR leaders discussed patient participation challenges, including cultural factors, financial challenges, accessibility issues, geographic location, health-related barriers, and system support barriers.

### Cultural and communication barriers

Participants described language differences, low health literacy, and culturally shaped perceptions of illness as barriers to participation, even when interpreter services were available. ^18–20^. For example, cultural attitudes toward health conditions can create barriers to blood pressure management, where patients may accept high readings as inevitable.

- *“We have a language line video interpreter phone everywhere in the hospital. There are small amounts on the total overall data of pockets of patients where English is not their first language”* (P1).
- *“We’ve thought a lot about how to increase referrals, especially from the Hispanic and Black population”* (P5).
- *“But I would say part of it is the culture…well, my blood pressure is high; it is what it is”* (P7).
- *“Literacy level is pretty low for our patients”* (P8).

### Financial and accessibility barriers

Patients and programs faced financial challenges, including insurance issues, high out-of-pocket costs, and conflicts with work responsibilities. For programs, financial constraints influenced staffing and resource limitations, resulting in understaffing and an expanded scope of practice for nurses ^13,21,22^. Accessibility issues involved transportation difficulties, high costs, and long waitlists that limit patient access to CR programs ^22,23^. Shared spaces and geographic constraints further limit participation, particularly for those in rural or underserved areas ^24–28^.

- *“We’ve seen patients with as much as a hundred-dollar co-pay per session. Patients often must choose between coming here and paying for medicine or food.”* (P5).
- *“Only three of us are on the team, so we have a significant waitlist”* (P5).
- *“Financial burden of going through all 36 sessions”* (P8).

### Health-related barriers

Health-related barriers included patient comorbidities, physical limitations such as muscle and joint pain, and functional impairments that restricted participation ^13,29^. Thus, tailored exercise interventions are necessary, yet some patients experience frustration or embarrassment due to mobility challenges ^5,13^.

- *“Patients say, ‘If I sit down in a chair, I can’t get back up.’ It’s frustrating and embarrassing for some patients.” “Patients feel frustrated or embarrassed about their physical challenges”* (P5).
- *“Comorbidities can make participation in rehab harder. Some patients just don’t have the energy or motivation especially older patients, heart failure patients, patients with physical barriers like joint and muscle pain”* (P9).
- *“Orthopedic issues like knee or back pain are common barriers”* (P10).

### System-level barriers

Additionally, the system faces barriers; including the lack of automated referral processes, which increase inefficiencies, and manual follow-up processes, which increase workload and impact on the workflow^13,30^. The COVID-19 pandemic exacerbated financial struggles for the healthcare system, and there was a need for updated medical equipment to improve patient care^31,32^.

- *“We don’t even have automatic orders yet at discharge.” “I have to go in and generate reports manually for cardiac follow-up”* (P2).
- *“Since the pandemic, it’s been very difficult financially”* (P7).

Despite these barriers, CR programs demonstrated multiple strategies aimed at improving participation, adherence, and quality of care, including policy changes, financial assistance programs, technological upgrades, and culturally tailored interventions to enhance participation and equity in CR ^5,13,28^.

### Strategies for Enhancing Accessibility and Outcomes

Programs emphasized the value of a patient-centered, adaptable, and evidence-based approach in CR programs. They combine clinical expertise, structured education, accessibility initiatives, and mental health integration with the goal to optimize patient outcomes and enhance program sustainability ^5,33^.

### Patient-centered and individualized care

Programs emphasized tailoring care to patient needs through adaptive exercise plans and individualized goal setting.

- *“We do mutual goal setting…at the end of your 36 sessions, you should be able to do XYZ.”* (P1).
- *“The best possible care we can. Individualize it”* (P3).

Moreover, mutual goal setting and risk factor modification have been highlighted as important for patient engagement (2,9). Leaders also recognized that small lifestyle changes, such as dietary improvements, could lead to significant health benefits. However, they also noted challenges, such as patient misconceptions about nutrition and varying motivation levels. Addressing these challenges requires personalized education and structured support to encourage long-term adherence (9,28).

- *“We focus on nine different risk areas to reduce risk.” “Each little change, like reducing sodium or increasing vegetables, can have huge health implications”* (P2).
- *“We’ve incorporated individual and group education, such as informal sessions with our dietitian, focusing on risk factor modification. However, we ensure that phase 2 patients receive focused education and that the staff feels confident using available resources. Regular quarterly check-ins and our close daily collaboration help strengthen this process”* (P4).

### Staff training and interdisciplinary collaboration

The results also highlighted key practices that influenced the success of CR programs including staff expertise, accreditation, individualized care, and patient-centered engagement. CR leaders emphasized ensuring all staff members are AACVPR-certified and undergo continuous training, including quarterly case reviews and competency assessments, to maintain high-quality care. The collaborative approach between CR staff members and cardiologists further strengthens the program’s credibility and effectiveness ^34,35^.

- *“All staff must be AACVPR certified, and new hires need to complete accreditation within a year”* (P1).
- *“I think that’s one of the biggest things we try to get the cardiologist and the thoracic surgeons on board with just sort of developing a report with us so that we could talk with them. And just making them aware of the importance of patients being here”* (P5).

### Accessibility-focused adaptations

Accessibility emerged as a critical factor, with leaders acknowledging that transportation barriers limit program participation ^11,28,36^. Leaders suggested strategies such as expanding community outreach and fostering partnerships with local organizations to improve awareness and inclusion, including free bus passes. Additionally, diversity in staff and patient populations was seen as a key factor in fostering an inclusive environment ^28,34,37^. Leaders also highlighted adaptations within CR programs, such as digitalizing individualized treatment plans to streamline care delivery and improve patient tracking ^2,8,38^.

- *“We expect they exercise at least 2 times outside of cardiac rehab on their non-cardiac rehab days. We also offer a free bus pass to the local fitness center here”* (P6).
- *“If they can’t come here and we can’t address those situations, we refer them somewhere else where they can get those things addressed as part of best practices.” We also have an inpatient cardiac rehab program in the hospital. They locate rehab programs for patients before discharge, ensuring a referral is sent to their local facility”* (P5).
- *“Diversity does help improve inclusion for everyone”* (P3).
- *“We have been going through the growing pains of adding a new EMR and transferring our original, individualized treatment plans or ITPs for the last year and a half”* (P3).
- *“We encourage patients right from the start by explaining that this program is designed to help them develop a consistent exercise habit while ensuring they do so safely. To support this, we provide exercise logs, typically within the first month of starting the program. Patients are given two-week increments of logs, which they complete and return to us. These logs help track their home exercise routines, and we use the information to guide their progress from there”* (P8).

### Multidisciplinary and psychosocial support

Moreover, access to a multidisciplinary team such as a cardiologist, a dietitian, dietitian, and a psychologist enriches patient support by addressing holistic needs, especially diet and mental health, which are integral to successful cardiac rehabilitation. Cardiac rehab programs need initiative-taking outreach to increase accessibility.

- *“If they have a disturbing rhythm during exercise, our cardiologists are on-site, making it easy to consult in real time”* (P1).
- *“Our dietician speaks Italian fluently, which helps with the Russian and Albanian population”* (P1).
- *“We have access to our social workers…should we feel like we need critical consultation”* (P4).
- *“We refer patients struggling with mental health to Dr. X, who has also been a patient in our program. Patients find his shared experience reassuring.”* (P5).
- *“Our system is called Health Partners…now we have many cardiologists coming to these outreach sites…We have much better access to cardiology and medical directors of cardiologists now, so we have input from them quite a bit”* (P6).
- *“We also have a referral to the pharmacist who is the smoking specialist”* (P6).

### Technology, virtual care, and engagement strategies

Virtual rehabilitation was identified as a potential future avenue, though insurance coverage remains a limitation ^39^. Recent federal policy extensions have supported the continued use of telehealth and remote rehabilitation services, though reimbursement and coverage remain variable ^14^. The integration of mental health support, including depression screening with the PHQ-9 and targeted interventions like journaling and webinars, underscores the growing recognition of psychological well-being in inpatient recovery ^40–43^

- *“We’re looking to potentially prepare for virtual rehab…not covered by most insurance, but we hope it will be in the future”* (P8).
- *“Hybrid-based cardiac rehab improved enrollment and attendance”* (P9).
- *“We use journals and rewards to encourage participation” one week we’ll do coloring. Talk about coloring. You know, things like that. That’s getting a gratitude journal.” “I have a bulletin board. And we have them in color. Let me tell them that when you’re finished, we will display it if you want, and they love that, like all this stuff.” “If you lose a pound, get a star; if you eat fish twice a week, you get a star”* (P7).*\*

### Continuity of care and referral systems

Furthermore, leaders supported the importance of continuity in care through structured referral processes ^9,26,44^. Efforts to ensure patients transition smoothly from inpatient to outpatient rehabilitation and receive appropriate follow-up care were critical in improving long-term outcomes ^9,45,46^.

- *“We work closely with cardiologists and primary care physicians, sharing detailed patient trends, medications, and outcomes”* (P2).
- *“Motivational interviewing is a big part of it. We are a small program compared to others, but each of us brings a unique personality and experience. Our different styles complement each other as we interact with patients”* (P4).
- *“We encourage patients to adopt exercise habits safely, using logs to track home exercises and progress”* (P8).
- *“We ask, do you have transportation to us? If they don’t, we offer some…a volunteer group or Medicaid services for transportation.” “Maybe you can’t make it three days a week. Sure, we’ll do two days…we try to be as flexible as we can”* (P10).

## Discussion

### Overview of findings

This study identified continuous multi-level barriers to CR participation alongside adaptive, patient-centered strategies used by high-performing programs. Together, findings highlight the complex interaction between structural, clinical, and social determinants influencing CR utilization and outcomes such as participation, adherence, and patient-centered care from the perspective of CR leaders.

### Barriers and equity implications

Findings showed that barriers to CR participation function at multiple levels; patient, program, and system, and disproportionately affect underserved populations. Cultural and language barriers persist even in programs with interpreter services, suggesting that structural resources alone are not enough without culturally responsive care models (18,20). Financial burden and transportation challenges further reinforce inequities in access, particularly among low-income and rural populations ^13,21,22^.

Health-related limitations and comorbidities also contribute to reduced participation, highlighting the need for adaptable and individualized rehabilitation approaches ^13,29^. System-level inefficiencies, including manual referral processes and staffing limitations, further compound access challenges and reduce program capacity ^13,30–32^.

### Interpretation of strategies

Programs responded to these barriers through a combination of individualized care, multidisciplinary collaboration, and flexible service delivery. These findings align with evidence that patient-centered CR models improve adherence and outcomes ^5,9^. Staff certification, interdisciplinary collaboration, and structured goal setting emerged as key mechanisms supporting program quality and consistency ^28,34,35^. Importantly, accessibility-focused adaptations such as transportation support and flexible scheduling reflect efforts to reduce structural barriers, though these remain inconsistently implemented ^11,28,34,36,37^.

### Innovation and future directions

Emerging strategies, including hybrid and virtual CR models, represent promising avenues for expanding access, although reimbursement limitations remain a barrier. The integration of mental health services and behavioral engagement strategies reflects a growing recognition of psychosocial needs in CR populations ^14,39^. System-level innovations such as automated referral systems and digital tracking tools may reduce administrative burden and improve continuity of care, but require further evaluation with an equity lens ^9,26,44–46^.

### Turning Evidence into Cardiac Care Equity

As an initial study exploring best practices in high-performing programs, these findings provide early insights into how CR delivery can be optimized to improve outcomes and reduce disparities. The implications of CR in education, policy, and research underscore the importance of comprehensive, evidence-based strategies for preventing and managing CVD. By working together to promote education, inform policy decisions, and advance research, healthcare providers, policymakers, and researchers can enhance outcomes for individuals with CVD and mitigate the overall public health burden ^5,16^. Collaborative efforts among healthcare providers, policymakers, and researchers to promote education, inform policy, and advance research can improve outcomes for individuals with CVD and reduce the overall public health burden ^11,14,44^.

### Next Generation Research Agenda

Building on this initial work, future research should prioritize equity and acceptability by prioritizing strategies that address key social determinants of health, including income, language, and geographic access. Expanding evaluation of hybrid and virtual CR models may help reduce disparities for rural and underserved populations facing transportation and access barriers particularly in light of recent federal policy extensions supporting telehealth and remote rehabilitation delivery, despite ongoing variability in reimbursement and coverage ^2,14,39,45^. Additionally, culturally tailored interventions are needed to address linguistic and cultural barriers that limit engagement and participation. ^9,14,19^.

Integrating mental health services into CR is another critical priority, given the unequal burden of depression and social isolation among low-income and underserved populations^23,24,41^. Finally, system-level innovations, such as automated referrals and enhanced use of patient tracking and care coordination systems should be evaluated with an explicit focus on equity and accessibility. These approaches may reduce administrative barriers and improve access for patients with lower health literacy, ensuring more timely and equitable participation in CR programs ^8,12,17,39,47^.

### Conclusion

This study identified multi-level barriers to CR participation, including cultural and linguistic differences, financial constraints, limited accessibility, patient comorbidities, and system-level inefficiencies in referral and documentation. While these challenges are well known, this study adds insight by showing how high-performing CR programs actively respond through adaptive, patient-centered strategies. These include individualized care, interdisciplinary collaboration, mental health integration, continuity of care practices, digital tools, and access-enhancing supports such as transportation and flexible scheduling. However, many strategies function as adaptive interventions rather than permanent solutions. Continued improvement in CR equity will require addressing challenging system-level barriers, including reimbursement, workforce capacity, and referral infrastructure.

## Data Availability Statement

The data that support the findings of this study are available from the corresponding author upon reasonable request.

## Authors’ Contributions

This integrative literature review was designed and written by R.T. (Raghad Tawalbeh). J.E. (Jullie Ellis), K.E. (Kyle T. Ebersole), and K.L. (Kim Litwack) served as committee members, contributing to the development and refinement of the review. R.T. was primarily responsible for manuscript drafting, with contributions from all authors to revision and interpretation of the literature. All authors contributed to critical revision of the manuscript for important intellectual content and approved the final version.

## Ethics Approval / Consent

Online verbal consent form

## Acknowledgements

We would like to sincerely thank the American Association of Cardiovascular and Pulmonary Rehabilitation (AACVPR) for their support in facilitating this study through access to program performance data and assistance with participant recruitment. We are especially grateful to the cardiac rehabilitation leaders who participated in this study for generously sharing their time, experiences, and insights. Their perspectives were essential in advancing understanding of strategies to improve cardiac rehabilitation accessibility, delivery, and patient outcomes.

